# “Early Imaging Marker of Progressive Glioblastoma: *a window of opportunity”*

**DOI:** 10.1101/2020.04.25.20079665

**Authors:** Na Tosha N. Gatson

**Affiliations:** Neuroscience Institute, Geisinger Health, Danville, PA, 17822; Cancer Institute, Geisinger Health, Danville, PA, 17822; Geisinger Commonwealth School of Medicine, Geisinger Commonwealth School of Medicine, Scranton, PA, 18509

**Keywords:** FLAIR signal intensity (SI), imaging biomarker, Neurologic Assessment in Neuro-Oncology (NANO), progressed glioblastoma, Response Assessment in Neuro-Oncology (RANO)

## Abstract

**Background:** Therapeutic intervention at glioblastoma (GBM) progression, as defined by current assessment criteria, is arguably *too late* as second-line therapies fail to extend survival. Still, most GBM trials target recurrent disease. We propose integration of a novel imaging biomarker to more confidently and promptly define progression and propose a critical timepoint for earlier intervention to extend therapeutic exposure.

**Patients/Methods:** A retrospective review of 622 GBM patients between 2006-2019 yielded 135 meeting resection, clinical, and imaging inclusion criteria. We qualitatively and quantitatively analyzed 2000+ sequential brain MRIs (initial diagnosis to first progression) for development of T2 FLAIR signal intensity (SI) within the resection cavity (RC) compared to the ventricles (V) for quantitative inter-image normalization. PFS and OS were evaluated using Kaplan-Meier curves stratified by SI. Specificity and sensitivity were determined using a 2×2 table and pathology confirmation at progression. Multivariate analysis evaluated SI effect on the hazard rate for death after adjusting for established prognostic covariates. Recursive partitioning determined successive quantifiers and cutoffs associated with outcomes. Neurological deficits correlated with SI.

**Results:** Seventy-five percent of patients developed SI on average 3.4 months before RANO-assessed progression with 84% sensitivity. SI-positivity portended neurological decline and significantly poorer outcomes for PFS (median, 10 vs. 15 months) and OS (median, 20 vs. 29 months) compared to SI-negative. RC/V ratio ≥4 was the most significant prognostic indicator of death.

**Conclusions:** Implications of these data are far-reaching, potentially shifting paradigms for glioma treatment response assessment, altering timepoints for salvage therapeutic intervention, and reshaping glioma clinical trial design.

**KEYPOINTS:**

- Increased confidence in defining true tumor progression is of critical importance.
- Imaging markers preceding progression offer novel timepoints for salvage therapies.
- Earlier intervention might increase tumor therapy exposure and reshape clinical trial design.

**IMPORTANCE OF STUDY:** Therapeutic intervention at progression has failed to show benefit. Accurately defining progression impacts clinical decision-making, yet current response assessment criteria in glioblastoma remain unvalidated. The data presented identifies a highly sensitive brain tumor imaging biomarker, SI, which coincides with declining neurologic function and might supplement existing criteria to improve clinician confidence to declare GBM progression. Furthermore, as SI precedes current assessment guidelines by an average of 3.4 months, this finding might also offer an earlier window of opportunity for salvage therapeutic intervention and reshape glioma clinical trial design. This signal has been previously associated with glioma progression; however, prior studies were hampered by overly inclusive criteria and failed to make the innovative clinical and prognostic associations evidenced in our study. Prospective validation of the proposed imaging biomarker is currently underway as part of a centrally reviewed prospective interventional clinical trial for newly diagnosed GBM.

## INTRODUCTION

Glioblastoma (GBM) is the deadliest primary brain tumor in adults, with median survival around 15 months despite aggressive upfront standard of care (SOC) treatment including maximal surgical resection followed by concurrent chemoradiation therapy and adjuvant temozolomide ^1,2^. GBM progression is near-universal and usually occurs at a median of around nine months; this is often followed closely by second progression within approximately 10-weeks ^3-5^. Despite its debated definition, recurrent/progressive GBM is associated with augmented tumor oncogenicity that renders second-line therapies ineffective, and there is no SOC for progressive GBM ^6-8^. Salvage therapies potentially fail due to poorer patient clinical tolerance at progression, rapid death after progression, and restricted therapeutic access to the central nervous system (CNS) depriving patients of adequate opportunity to expose the tumor to sufficient drug ^6,7,9-13^. Therapeutic resistance after progression has also been attributed, in part, to oncogenic phenocopying, enrichment of resistant glioma stem cells, immunomodulation, increased tumor heterogeneity or mutational burden, and delays recognition and start to therapy ^11,14-18^.

Defining tumor progression is an active topic in neuro-oncology as it applies to clinical practice and standardization of clinical trial imaging technique. Nonetheless, current guidelines defining progression remain unvalidated ^19-24^. There continue to be limits to consensus in discerning true tumor progression from treatment-related changes or pseudoprogression on brain imaging at the time of declared radiographic progression ^21,24-26^. Inadequacies in earlier identification of tumor progression adversely impact clinical decision-making for effective GBM salvage treatment.

In 1990, Macdonald *et al*. published criteria for response assessment in high-grade glioma which assessed tumor volume using 2-dimensional imaging criteria ^19,20^. However, this only addressed the contrast-enhancing component of the tumor and failed to consider clinical factors such as corticosteroid use and neurologic status ^20,22^. In systemic cancers use of a one-dimensional tumor measurement protocol is detailed in the updated Response Evaluation Criteria in Solid Tumors (RECIST v1.1) which determines progress as an increase in longest tumor diameter of at least 20% from baseline imaging ^22,23^. Studies suggest good concordance of RECIST with 2-dimensional criteria ^22,27^, however, RECIST has not been prospectively validated in high grade glioma. Due to the limitations of the Macdonald Criteria, the efforts in neuro-oncology to improve imaging response assessment in high-grade glioma and standardization of imaging for clinical trials led to the development of the Response Assessment in Neuro-Oncology (RANO) Working Group ^21,22,27^. The RANO criteria combine two-dimensional tumor measurements on computed tomography or MR imaging, account for both the enhancing and non-enhancing tumor and considers patient clinical assessment and corticosteroid use ^21,27^. While RANO has become the mainstay of assessment in glioma treatment response, this too remains unvalidated. In the future, evolving volumetric and physiologic imaging techniques might be validated as response tools. In the meantime, standard MRI might harbor hitherto unexplored early radiographic indicators of progression. Assessment of such features would be useful to support decision-making when there are equivocal clinical or conventional imaging findings for tumor progression. One such potential feature is a change in MRI T2-weighted fluid attenuated inversion recovery (FLAIR) signal hyper-intensity in the surgical resection cavity.

The resection cavity is typically isointense to cerebrospinal fluid (CSF) on FLAIR MR imaging. Winterstein *et al*. (2010) retrospectively evaluated FLAIR MRI findings of 75 subjects including all glioma grades (World Health Organization [WHO] grades I-IV), partially resected gliomas and radiation therapy in select patients. The “standard call” for progression was made using RECIST criteria. However, when they utilized FLAIR signal intensity as an indicator of first progression (which almost universally occurred prior to RECIST-designated progression), they had 100% specificity and 57% sensitivity for progression. Their group was the first to propose the use of this approach to designate progression and postulated that the signal increase in the resection cavity is a manifestation of early encapsulation of the resection cavity by tumor cells ^28^. A later study by Ito-Yamashita *et al*. (2013) retrospectively evaluated 44 subjects, also with partially resected high-grade gliomas (WHO grades III-IV) after radiation therapy. FLAIR signal increase within the resection cavity prior to or at the time of RECIST-designated progression has a 100% specificity for disease progression but lower sensitivity (34%) as compared to the initial study ^29^. Importantly, their group determined that this technique has higher sensitivity for estimating progression in WHO grade IV gliomas compared to WHO grade III gliomas ^29^. More contemporary studies, done by Sarbu *et al*. (2016) and Bette *et al*. (2017), used RANO assessment criteria and evaluated WHO grades II-IV for FLAIR changes within the resection cavity. The study by Sarbu *et al*. included gross totally resected (GTR) patients and demonstrated improved sensitivity to 65%, whereas the study by Bette *et al*. included patients with subtotally resected or biopsied tumors, resulting in a lower sensitivity (18%) ^30,31^.

Earlier detection of radiographic disease progression could lead to improved clinical decision-making, and earlier utilization of therapies could potentially enhance their efficacy and improve patient outcomes. Our study uses more stringent inclusion and exclusion criteria and applies novel integration of clinical and tumor molecular features in the assessment of FLAIR signal hyperintensity (SI) within the resection cavity prior to progression. These findings could serve as harbingers of progression and potentially supplement current response assessment criteria. Furthermore, results of the ongoing prospective validation study will be helpful to establish whether this imaging biomarker provides a viable earlier timepoint for therapeutic intervention.

## METHODS

### Study Objective and Design

This was a noninterventional, large, single institution retrospective review of patients diagnosed with WHO grade IV astrocytoma, initiated on SOC therapy between 2006-2019. Over 2000 baseline and follow-up MR imaging studies prior to the first RANO-criteria radiographic progression were reviewed. With strict inclusion and exclusion criteria, we analyzed radiographic, clinical, and pathomolecular data using both qualitative and quantitative techniques to identify early indicators of progression. We explored the impact of SI on progression free survival (PFS) and overall survival (OS) on a subset of cases between 2016-2019 adjusted for O^6^-methylguanin-DNA-methyltransferase (MGMT) status and analyzed the association of elevated monoclonal antibody proliferation marker index (MIB-1) with risk of SI within the resection cavity. This study was approved by the Geisinger Health Institutional Review Board (IRB, #2018-0274) in accordance with the standardized ethical principles in relation to human subject’s research and patient confidentiality.

### Patient Population

This is a retrospective review of 622 adults (≥18-years) with histopathologically confirmed, newly diagnosed glioblastoma or gliosarcoma treated between January 1, 2006 – September 1, 2019. Of these, 487 were excluded from analysis as shown in **Figure 1**. Briefly, we excluded 263 patients with insufficient imaging and clinical data or less than 10-months follow-up. Another 162 patients with collapsed resection cavity, biopsy/subtotal tumor resection, or resections involving the ventricles were unevaluable and hence excluded. An additional 58 were excluded because of positive isocitrate dehydrogenase (IDH)-mutation; this was done to enhance molecular homogeneity of the final population evaluated in our study. Four patients with distal progression outside of the original resection cavity were also excluded. This allowed us to evaluate a total of 135 patients.

**Figure 1.**
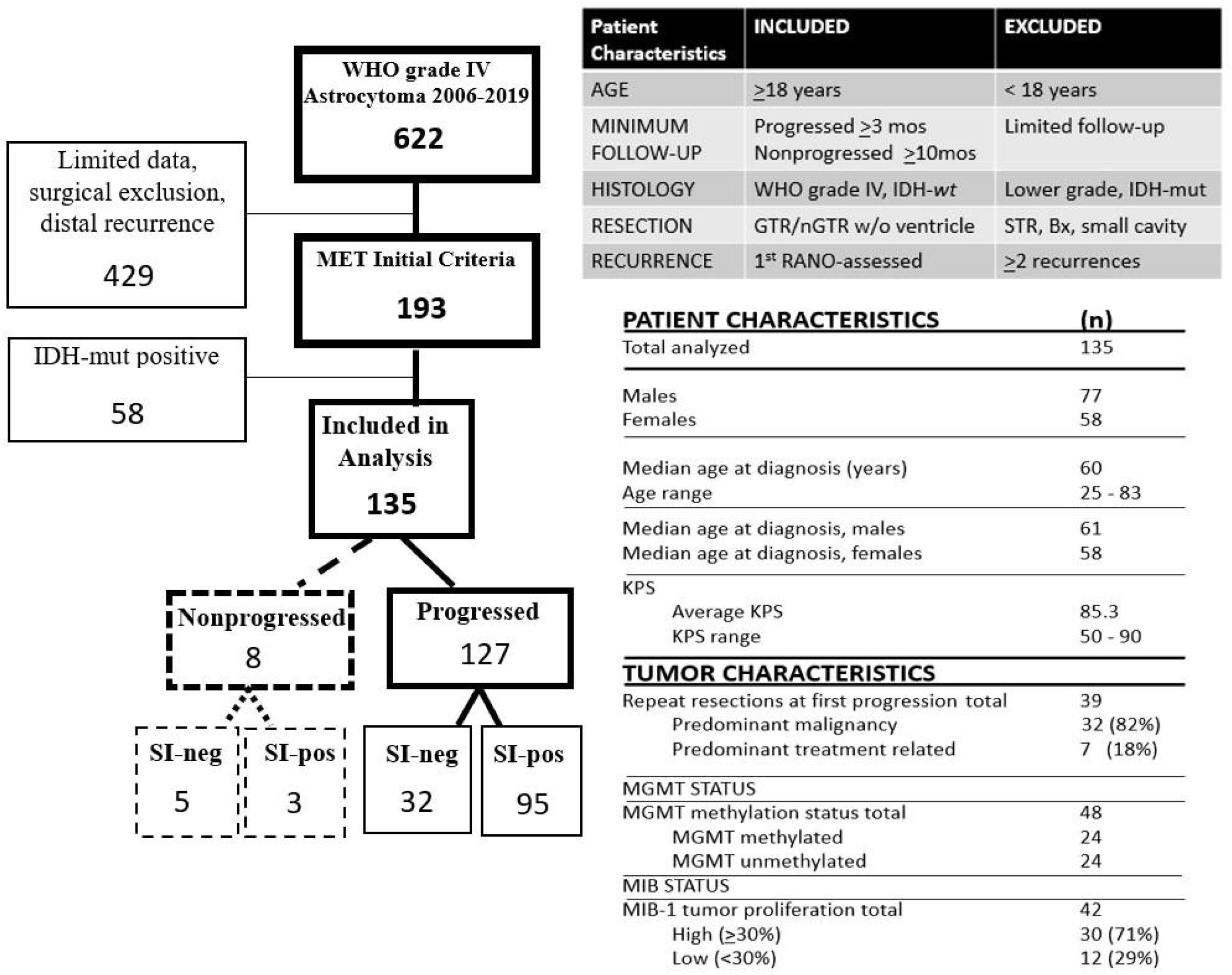
Intent to Evaluate Tree **(left)**. Inclusion and exclusion criteria **(right top)**. Patient and tumor characteristics **(right bottom)**. *Signal Intensity* (*SI*); *IDH mutated* (*IDHmut*); *Negative* (*neg*); *Positive* (*pos*)

GTR or near gross total resection (nGTR, ≥90% of contrast enhancing tumor) was confirmed using post-operative brain MRI within 72 hours. Patients were followed through the first declared radiographic progression in accordance to the RANO criteria ^21,32^. All 135 cases included for final analysis were identified by progression status as progressed (P) or nonprogressed (NP). They were also dichotomized, by presence or absence of FLAIR SI within the resection cavity, as either SI positive (SI-pos) or SI negative (SI-neg). Patient characteristic and demographics are summarized in **Figure 1**.

### Primary and Secondary Endpoints

Date of diagnosis was defined as date of initial surgical resection. Time to first progression (TTP) was calculated from date of diagnosis and represented the progression free survival period for the purposes of this study. Study primary endpoints were PFS and OS. The secondary endpoints evaluated relationships between SI and age, Karnofsky Performance Scale (KPS), and sex, and the possible impact of these associations on the primary outcomes.

### Exploratory Analysis

In order to determine whether MGMT or MIB-1/Ki-67 status had any association with SI, we performed an exploratory analysis on a smaller patient subset (selected from 2016 and 2019, when these assay results became routinely available at our institution). These preliminary data estimated the impact of SI on PFS and OS in relationship to tumor MGMT status (n=48) and MIB-1/Ki-67 proliferation index (n=42).

### Neuropathology and Tumor Molecular Confirmation

Histopathological designation of WHO grade IV astrocytoma/glioblastoma or gliosarcoma was assigned based on the 2007 and 2016 WHO criteria for CNS tumors ^33,34^. Immunohistochemical stains were performed on formalin-fixed and paraffin-embedded 4-*μ*m routine tissue sections. Standard, previously defined molecular techniques for IDH1 R132H, p53, Ki 67, ATRX, Olig2, H3K27M analysis, using appropriate antibodies on deparaffinated tissue were employed. Paraffin blocks were forwarded to an outside lab (NeoGenomics) to test for epidermal growth factor receptor amplification (EGFR) by fluorescent in-situ hybridization (FISH), and MGMT Gene Promotor Methylation and IDH 1 and 2 mutations analysis by polymerase chain reaction (PCR) for patients under age 55 or with need for further confirmation.

### Defining tumor progression

Radiographic tumor progression was defined using RANO criteria ^21^. For patients who underwent repeat resection at first progression (n=39), pathology report was reviewed for confirmation of tumor recurrence versus treatment related changes. These cases were then dichotomized based on their SI status as another approach for determining accuracy for predicting progression.

### MRI Protocol

MRIs were performed on either 1.5 or 3 Tesla MRIs using a pre-defined institutional tumor protocol. The majority used 2D T2-weighted FLAIR images in the axial plane, using 5-mm slice thickness with a 1-mm interslice gap; fewer than 10% of cases used 3D FLAIR. Although specific parameters varied across magnets, the use of reference internal controls allowed for comparison between scans. MRIs were collected within 72 hours post-operative and thereafter every 2-4 months after completion of chemoradiotherapy for evaluation.

### Image analysis and SI Assessment

Imaging was reviewed by a neuroradiologist with significant brain tumor imaging experience and by a practicing neuro-oncologist.

#### Qualitative analysis

Signal intensity was assessed within the resection cavity (RC) on FLAIR imaging and determined as hyperintense relative to the ventricles (V) in the same study. All subsequent MRIs obtained prior to first progression were reviewed and scored as hypointense, isointense, or hyperintense for the RC as compared to the V. SI was determined once there was confirmed qualitative change in FLAIR hyperintensity within the RC as compared to V but at least 3 months after resection to reduce hyperintensity error secondary to post-operative blood within the RC. Time to SI-pos signal (TTSI) was measured as time from diagnosis to development of hyper-intense signal within the RC. Early TTSI was defined as signal development < 5 months; Intermediate TTSI ≥5 but <11 months; and Late TTSI ≥11 months. Time to progression from onset of SI (TTSI-P) was measured as time between defined SI to RANO-assessed progression.

#### Quantitative analysis

Quantitative imaging analysis was performed as described in Winterstein *et al*. (2010), except for the modifications as described below. The RC, primary area of interest, and ventricles were measured independently using NIH ScionJ imaging software (ImageJ News Version 1.52t 30) to obtain objective values for signal intensity within the RC and V compartments at three timepoints:

- pre-SI MRI, (2) when qualitative SI-pos declaration was made, and (3) at the time of RANO-assessed progression. For the RC, three sample circles of equal area (minimum of 15mm × 15mm) were drawn within the RC and values for intensity of the signal were averaged. The modification using three smaller circles within the area of interest allowed increased accuracy of the RC measurement, facilitated measurements for variable resection cavity conformations and reduced the risk of including brain parenchyma in the selected region. Each of the measurements were averaged to calculate the value for RC in each patient using *intensity units*. For the ventricles, two circles within the ipsilateral ventricle (as compared to initial tumor location) and one contralateral circle were created, and measurements were averaged to calculate the intensity unit value for V. The inclusion of both ventricles minimized potential noise, bias, or variability related to ventricle proximity to the treated RC **(Figure 2)**.

**Figure 2.**
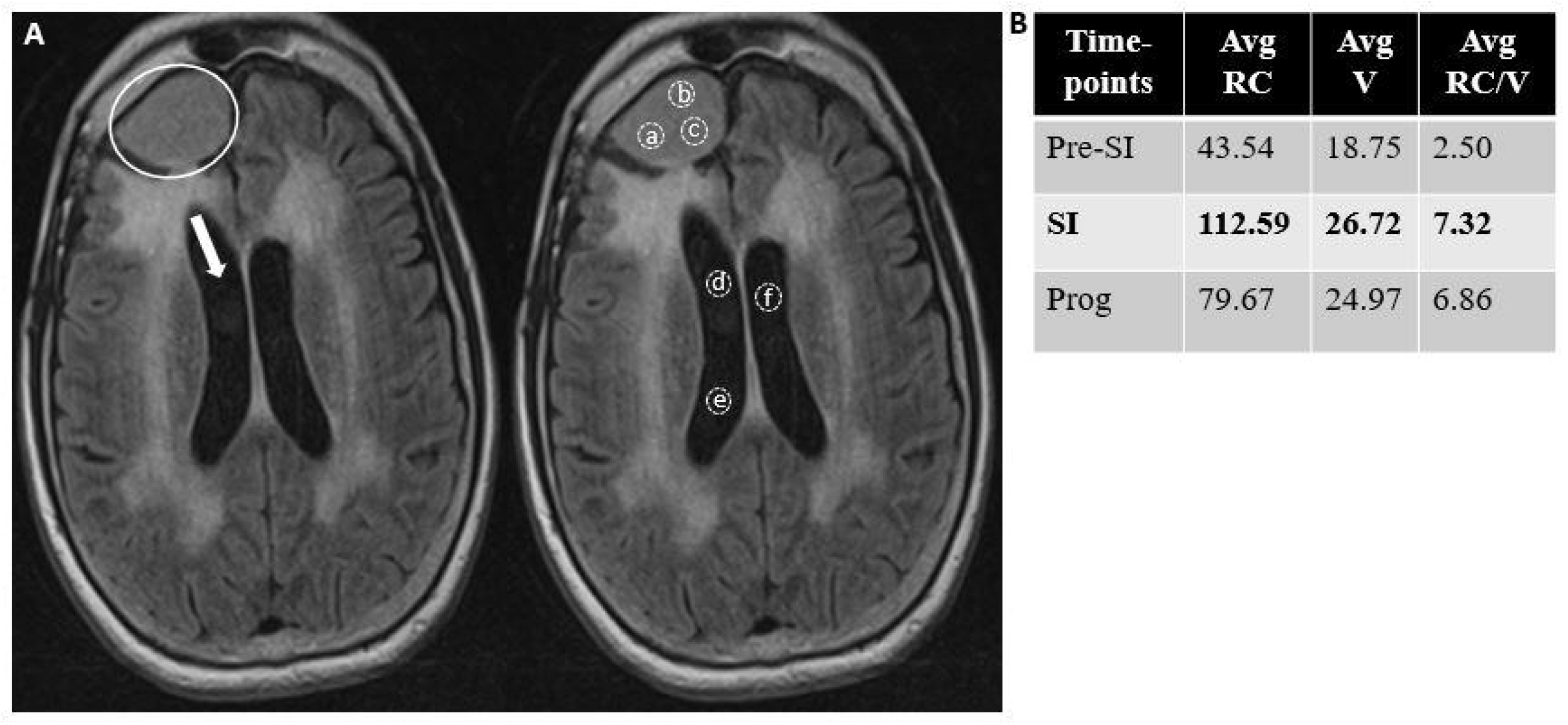
(A, Left image) Qualitative analysis (FLAIR) MR brain imaging illustrates increased SI within the RC (left image, white circle) as compared to V (left image, white arrow). (A, Right image) Quantitative analysis of brain MR imaging for FLAIR SI within RC (averaged 3 measurements *a,b,c*), and within the V (averaged two ipsilateral measurements *d,e*; and one contralateral measurement *f*). (B.) RC/V ratio measures for three timepoints: pre-SI, SI, and progression. *Progression* (*Prog*); *Resection cavity* (*RC*), *Ventricle* (*V*)

### NANO Scale Clinical Assessment and Relationship to SI

Review of the electronic medical record (EMR) documentation of clinical assessments at routine visits at time of brain MRI collection was performed to determine clinical score in accordance with Nayak *et al*. (2017) to assess neurological function for integration with RANO criteria [*Neurologic Assessment in Neuro-Oncology* (NANO)], at three timepoints: (1) pre-SI MRI, (2) when qualitative SI-pos declaration was made, and (3) at the time of RANO-assessed progression ^10^. Patients were assigned a NANO scale score based on assessment of nine relevant neurological domains measured as part of NANO. Scores were compared for significant differences at assessed timepoints, relative to SI.

### Statistical analysis

Primary outcomes PFS and OS were evaluated using Kaplan-Meier curves, stratified by SI. The log-rank test was used to assess the difference in survival curves between the SI groups for each of the primary outcomes. Secondary outcome was time to development of SI, stratified by age, sex, KPS, MGMT status, and MIB-1 index. Univariate Cox proportional hazards models were performed to assess the impact of SI on the hazard rate of primary outcomes. Multivariate Cox proportional hazards model assessed the effect of SI on the hazard rate for death, after adjusting for known prognostic confounding variables for survival including age, sex, and KPS. The Hazard Ratios (HRs) and corresponding 95% confidence intervals (CIs) were computed. A decision-tree-algorithm (recursive partitioning analysis) was implemented to represent decision-making in predicting the classification label: survival or not. Decision tree algorithm implicitly performed feature selection from input variables including age, sex, KPS, MGMT status, and RC/V ratio. Recursive partitioning analysis was used to determine which successive quantifiers or value cutoffs, specifically for RC/V ratio, sex, and age were most strongly associated with survival versus death. Variables not shown on the decision-tree did not demonstrate an impact on survival at a rate higher than those shown in the diagram. SI relationship to MGMT and PFS or OS were exploratory due to small sample size. We used a two-by-two table to assess the positive predictive value (PPV) and negative predictive value (NPV) and determined the sensitivity and specificity of the SI for median PFS-6/12 and median OS-6/12 for comparison to landmark data. We also used pathology confirmation on repeat resection for SI-pos/neg patients at time of declared progression as another measure of accuracy. Statistical analyses were performed in RStudio (Version 1.2.5019). P-values of less than 0.05 were considered statistically significant.

## RESULTS

### Summary of Evaluated Patient Population

Of the 135 eligible patients, 57% were males (n=77). Median age at diagnosis was 60 years [range 25-83]. Ninety-four percent (n=127) had RANO-assessed radiographic progression of which 75% (n=95) were SI-pos and 25% (n=32) were SI-neg. The median follow-up time was 19.3 months (range 10 to 166). By the end of the study, only 6% (n=8) were non-progressed, of these 38% (n=3) were SI-pos and 63% (n=5) were SI-neg. For the SI-pos group, the probability of progression at 6- and 12 months was 33% and 60% vs. 19% and 41% for the SI-neg group. The probability of death at 6- and 12 months was 3% and 82% for the SI-pos group vs. 0% and 65% for the SI-neg group (*see* ***Supplemental Figure S1A*)**. After RANO-assessed progression, 39 (31%) patients underwent repeat resection, of these 29 (74%) were SI-pos while 10 (26%) were SI-neg. Within the SI-pos group, 89% (n=26) had pathology confirmed recurrence vs. 60% (n=6) within the SI-neg group. This approach yielded a sensitivity and specificity of 84% and 71%, respectively, with a PPV of 93% and NPV of 50% (*see* ***Supplemental Figure S1B*)**.

The mean time from SI-pos signal to RANO-assessed progression was 3.4 months. The objective measure of SI-pos was RC/V ratio (Figure 2A). Figure 2B demonstrates the mean RC/V ratios at three timepoints: pre-SI (2.5), at SI (7.32), and at progression (6.86).

### Impact of SI and RC/V ratio on PFS and OS

For all included cases, the median PFS and OS were 10 months and 18 months, respectively. The SI-pos group had poorer outcomes as compared to the SI-neg group. The median PFS for the SI-pos vs. SI-neg groups were (10 vs. 15 months) [p=0.0037, HR 1.733, 95% CI 1.208-2.485]. The median OS for SI-pos vs. SI-neg groups was 20 vs. 29 months [p=0.0047, HR 1.871, 95%CI 1.254-2.793] **(Figure 3)**. Multivariate Cox proportional hazard model for OS indicated that 1.88 times as many SI-pos patients experienced death as compared to the SI-neg patients (HR = 1.88, 95% CI: 1.17 to 3.02, p =0.0087), while the PFS model indicated 2.45 times as many SI-pos patients experienced progression (HR=2.45, 95% CI: 1.50 to 4.00, p =0.00325) after adjusting for age, sex, and KPS **(*see Supplemental Figure S2*)**.

**Figure 3.**
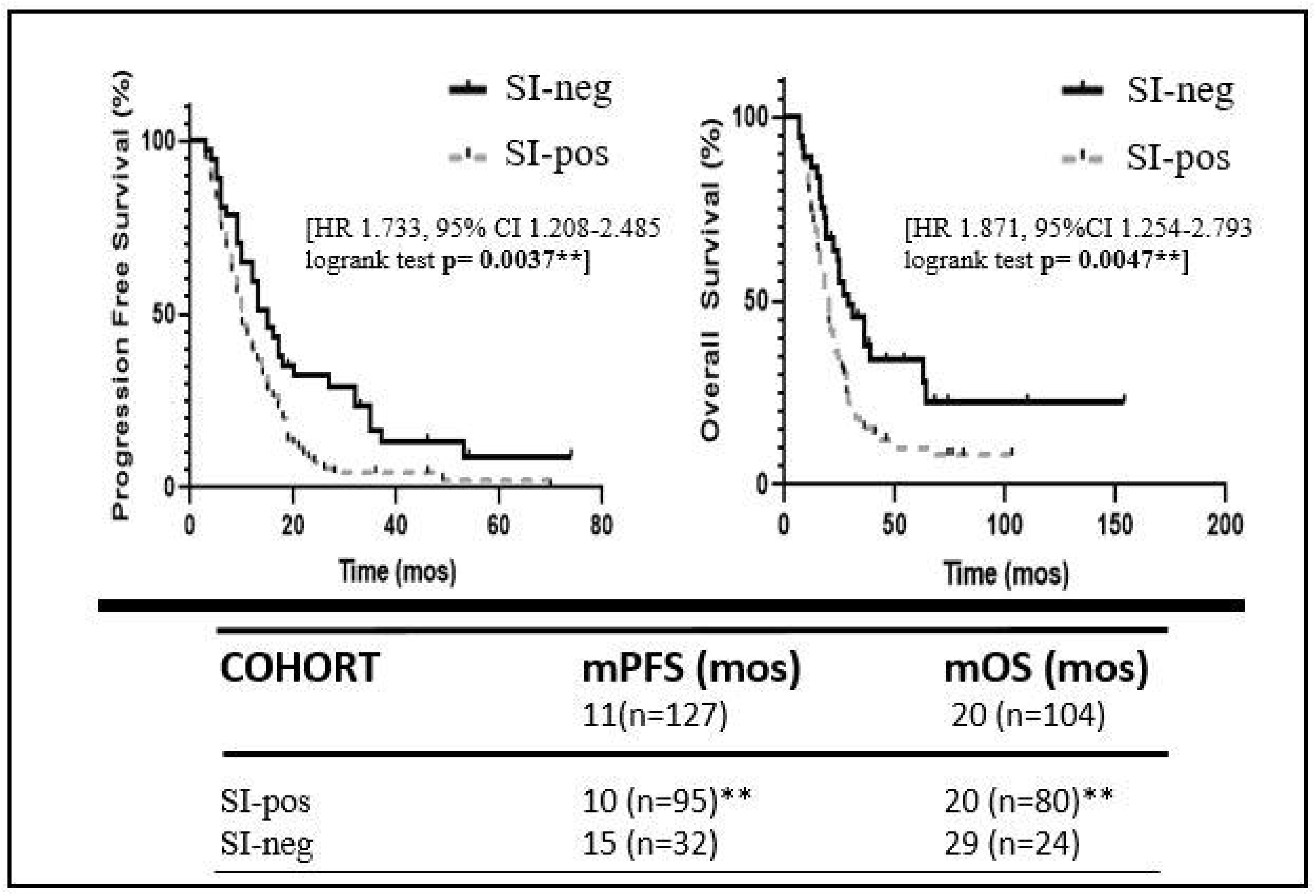
Analysis of SI Impact on Progression Free Survival and Overall Survival. The SI-pos group shorter PFS and OS as compared to SI-neg group. PFS **(Left K-M curve)** and Overall Survival **(Right K-M curve)**.

RC/V ratio ≥4 was determined by the algorithm as the first node in the decision tree for the binary outcome of survival vs. death. RC/V ratio ≥4, female sex, and age ≥64 were the combined variables with the highest risk for death **(*see Supplemental Figure S3A*)**. The RC/V ratio was inversely proportional to PFS and OS, most significant decline after RC/V ratio of ≥4. **(*see Supplemental Figure S3B*)**.

### Exploratory Survival Analysis on impact of MGMT status dichotomized by SI

MGMT methylated (M) cases (n=24) had longer median PFS and OS (15 and 28 months) as compared to MGMT unmethylated (U) cases (n=24) at (9 and 19 months). The median PFS and OS for the M-SI-pos group vs. M-SI-neg group was 14 and 28 vs. 16 and 64 months, respectively. The median PFS and OS for the U-SI-pos group vs. U-SI-neg group was 9 and 19 vs. 6 and 36 months, respectively. The M-SI-neg group demonstrates the longest mOS (64 months), whereas the U-SI-pos group demonstrated the shortest mOS (19 months).

As compared to the median PFS and OS for all cases within the MGMT-M and -U groups, there was no further impact when dichotomized based on SI-*pos* status. However, when dichotomized based on SI-*neg* status there was a notable impact on survival outcomes **(*see Supplemental Figure S4***). There was no reliable trend for survival outcomes for MIB-1, EGFR, or TP53 status when dichotomized based on SI-status in this exploratory group (data not shown).

### Secondary Outcomes Measures

#### NANO clinical assessment scale relationship to SI

NANO scale was significantly lower at the pre-SI timepoint vs. the SI timepoint (1.14 vs. 1.88; log rank test p=0.00002**). However, there was no significant difference between the NANO scale at the SI timepoint and at the time of progression (1.88 vs. 2.02; p=0.466) **(Figure 4)**.

**Figure 4.**
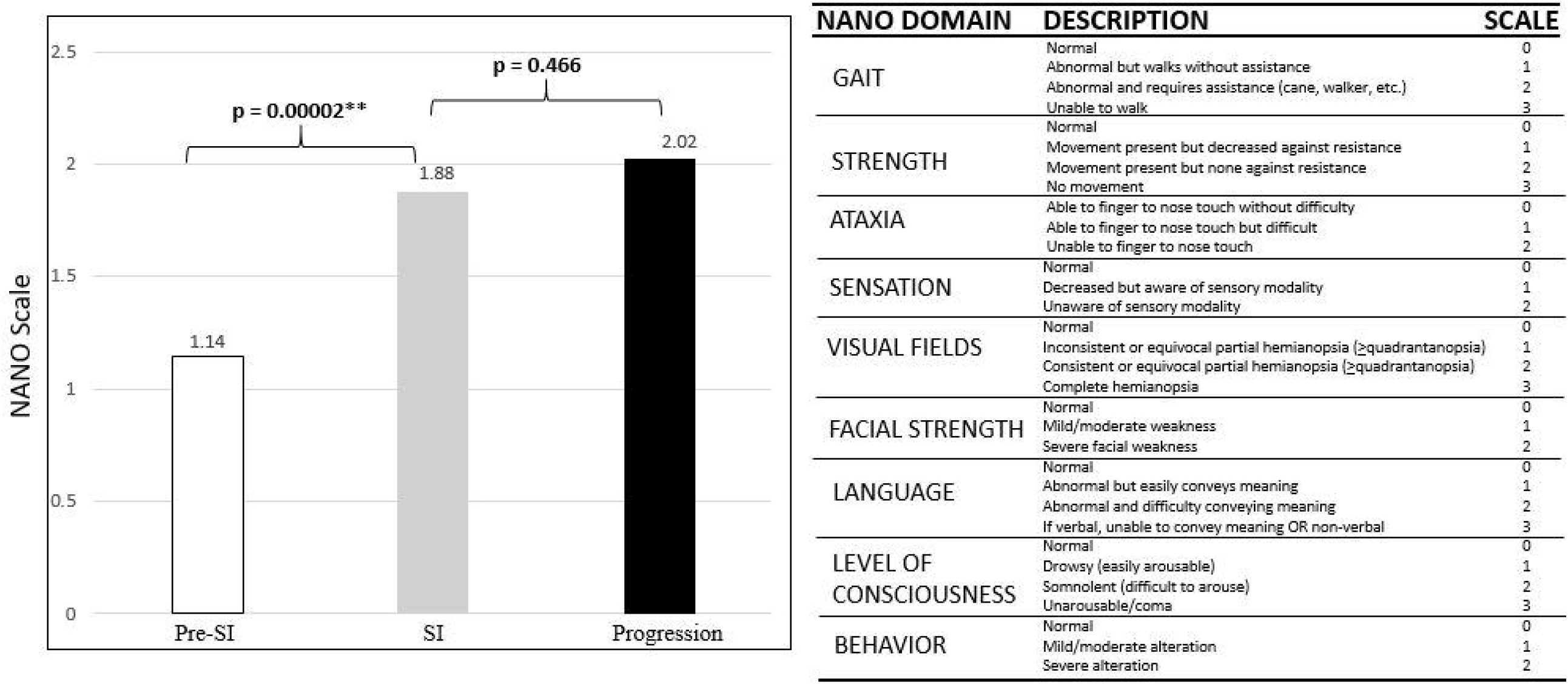
**(Left)** Comparative analysis of the NANO scale for all 135 patients at three timepoints, pre-SI, SI, and progression. The average NANO scale at SI and progression are both significantly higher than at pre-SI. **(Right)** NANO Scale Domain – Nayak *et al*. 2017. *Neurologic Assessment in Neuro-Oncology* (*NANO*)

#### Time to SI (TTSI) relationship to PFS and OS

Longer TTSI correlated with longer mPFS and mOS outcomes. For Early vs. Intermediate vs. Late TTSI mPFS was (6, 10, 19 months; p=0.0001****) and mOS was (12,18, 26; p= 0.0001****). (***see Supplemental Figure 5***).

**Figure 5.**
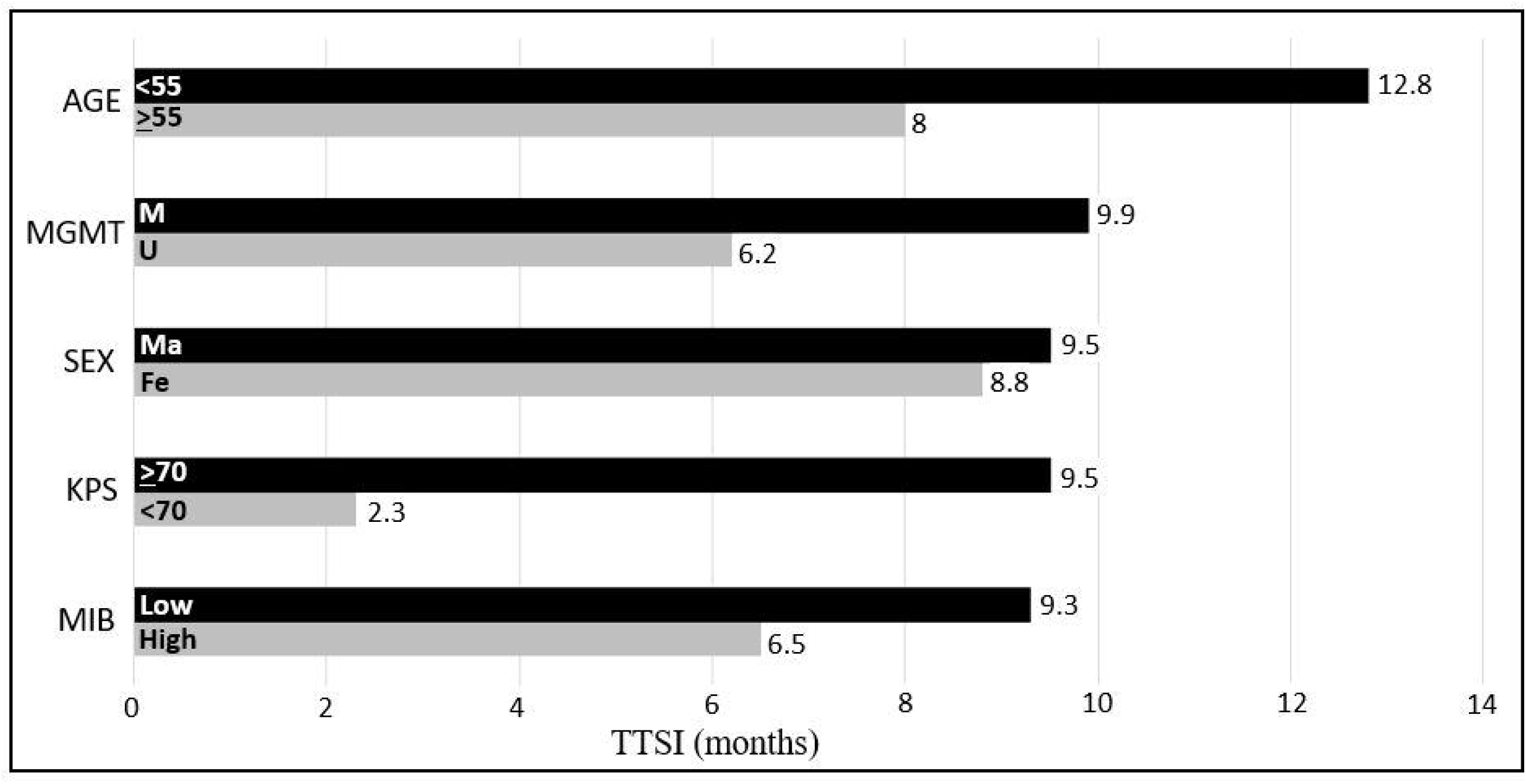
Univariate Analysis of Known Survival Variables Influence on Time To SI (TTSI). Patients who are younger with methylated tumors that have higher KPS and a lower MIB-1 are more likely to have delayed development of SI signal. No sex influence demonstrated. *Female* (*Fe*); *Karnofsky performance scale* (*KPS*); *Male* (*Ma*); *Methylated* (*M*); *Monoclonal antibody proliferation marker-1* (*MIB-1*) <*30%* (*low*); *MIB* ≥ *30%* (*high*); *O-6-methylguanine-DNA-methyltransferase* (*MGMT*); *Time to SI* (*TTSI*); *Unmethylated* (*U*)

#### Factors influencing TTSI

The average TTSI for all cases was 9.07 months. Shorter TTSI was observed in patients ≥55-years-old at diagnosis, MGMT unmethylated status, and higher MIB-1 indices (≥30%). KPS ≥70% demonstrated the longest TTSI (12.8 months) and KPS <70% demonstrated shortest TTSI (2.3 months). There was no significant sex-influence on TTSI **(Figure 5)**.

#### Factors influencing time interval between SI-pos and RANO-assessed progression (TTSI-P)

Average TTSI-P was 3.4 months. MGMT methylated tumors had a longer average TTSI-P (5.2 months) as compared to MGMT-unmethylated tumors (3.6 months) **(*see Supplemental Figure S6***). Interestingly, female patients and patients < 55 years old at diagnosis demonstrated a trend toward shorter TTSI-P (2.6 and 3.1 months) respectively. There was no significant impact on TTSI-P by MIB-1 index or KPS **(*see Supplemental Figure S7*)**.

#### Factors influencing the magnitude of the RC/V ratio at time of SI signal development

The mean RC/V ratio at time of SI was 7.32. A higher average RC/V ratio was observed in MGMT unmethylated tumors as compared to methylated (10.3 vs. 6), *data not shown*. Higher tumor MIB-1 index demonstrated increased average RC/V ratio as compared to low MIB-1 index tumors (12.2 vs. 4.8), *data not shown*.

## DISCUSSION

### Significance of the Study

This study uses routine brain MRI surveillance in high-grade glioma along with clinical and molecular pathology data to better predict tumor progression. Clinical trials designed to intervene at the time of RANO-assessed recurrence have failed to significantly improve overall survival – making this timepoint effectively *too late* ^3,22,23,35^. This is an active topic in neuro-oncology regarding patient care and standardization of imaging techniques with implications for clinical trial design ^32, 36-38^.

### Uniqueness of the Study

Stringent inclusion criteria of patients with the highest-grade astrocytoma and greatest extent of resection allowed for this first report from United States to provide evidence of a measurable imaging biomarker, *SI*, that precedes progression with a higher sensitivity than prior studies ^28-31^. We uniquely integrated clinical performance analyses and tumor molecular markers in association with SI to determine survival impact. A comparative review of this and prior studies is provided in **Table 1**.

**Table 1.** Comparative analysis of the included study (Gatson *et al*. 2020) as compared to the four prior reports of FLAIR signal anticipating progression. Prior studies had wider inclusion criteria in comparison. **\***Two-by-Two models to positive and negative predictive value to measure sensitivity and specificity of the SI as shown in *Supplemental Figure S1*.

| Study Year, Author<br>Country | WHO<br>grades<br>Included | Study<br>Sample<br>Size | Response<br>Assessment<br>Criteria<br>Utilized | Extent of<br>Resection<br>Included | Re-Resection at<br>1<sup>st</sup> Progression | Sensitivity<br>&<br>Specificity | Clinical<br>Score<br>Integration | Analysis of<br>Molecular<br>Markers |
| --- | --- | --- | --- | --- | --- | --- | --- | --- |
| 2010, Winterstein *et al.*<br>Germany | I – IV | 75 | RECIST | Partial only<br>*excludes GTR* | No Data | **57% Sensitivity**<br>100% Specificity | None | None |
| 2013, Ito *et al.*<br>Japan | III – IV | 44 | RECIST | Partial only<br>w/o ventricle | No Data | **34% Sensitivity**<br>100% Specificity | None | None |
| 2016, Sarbu *et al.*<br>Spain | II – IV | 107 | RANO | “Curative<br>intent” | No Data | **65% Sensitivity**<br>75% Specificity | None | None |
| 2017, Bette *et al.*<br>Germany | II – IV | 212 | RANO | All resection<br>types<br>evaluated | Yes | **18% Sensitivity**<br>80% Specificity | None | None |
| **2020, Gatson *et al.***<br>**USA (Current Study)** | **IV** | **135** | **RANO** | **GTR & Near GTR**<br>**-subgroup**<br>**ventricle**<br>**comparing (eye)** | **Yes**<br>**84% Sensitivity**<br>**71% Specificity**<br>**\*(Pathology**<br>**confirmation)** | **75% Sensitivity**<br>63% Specificity<br>**\*(Radiographic**<br>**confirmation)** | **KPS,**<br>**NANO**<br>**scale** | **IDH,**<br>**MGMT,**<br>**MIB-1,**<br>**EGFR, &**<br>**TP53** |

### Pathophysiology of SI

The pathophysiology of the development of SI within the RC is not well understood, but may correspond to cerebral spinal fluid (CSF) trapping, increased cellular expression of proteins within the resection cavity, and increase permeability of newly formed vessels in progressive tumors leading to leakage of proteins and blood products within the cavity ^28,31,39-41^. Two studies found that prior radiation exposure variably impacts the degree of SI ^28,31^.

Tumor cell encapsulation of the RC leading to CSF trapping and concentration of protein is a leading hypothesis for SI development ^28,29^. Surgical ventricle communication with the RC leads to reduced SI but limits ability to quantitate signal changes ^31^. We completed a unique subanalysis to quantify the FLAIR signal within the vitreous chambers of the globes (eye) on axial brain MRI as an alternative fluid attenuated cavity for comparison, noting comparable RC/Eye and RC/V ratios **(Figure S8)**.

We propose the expression of oncogenic proteins into the RC by surrounding glioma and glioma stem-like cells contributes to SI development. Further malignant differentiation of adjacent cells and changes in the tumor microenvironment possibly down-regulate this protein expression, leading to the observed fading of SI over time. MGMT activation and inactivation cycles are specific to the tumor microenvironment, including exposure to glucocorticoids, and might correlate with observed imaging changes ^42,43^. Accurate assessment of MGMT promoter methylation status and correlative protein expression are active topics in neuro-oncology ^44^. These and other studies designed to elucidate the biochemical and pathophysiological basis for SI are ongoing in our labs.

### Timing of Therapeutic Intervention in Gliomas

The survival impact of timing of therapeutic intervention in newly diagnosed gliomas has been variably addressed. Studies in low-grade gliomas have overall favored surgical intervention as opposed to the watch-and-wait approach ^45^. There is less consensus regarding the time to initiation of chemoradiotherapy in high-grade gliomas, but guidelines recommend initiating chemoradiation within 6 weeks of surgery; however, extent of resection and tumor molecular markers were not fully dichotomized ^46,47^. Metronomic use of systemic chemotherapy may hamper selective oncogenic tumor features, however, without improving overall survival ^48-51^. Therapeutic timing in the recurrent setting is limited by the ability to promptly and confidently identify progression. Regardless, therapies introduced at progression have been generally ineffective. Earlier intervention prior to radiographic progression might increase the duration of tumor cell exposure to therapeutic agents and allow for determining true clinical benefit of salvage therapies, and potentially delay neurologic functional decline. Earlier tumor targeting might also serve to decrease the tumor mutational burdens that render salvage therapies ineffective.

### Implications for Clinical Trial Design, a Window of Opportunity

We describe an identifiable imaging marker, after initiation on SOC, which reliably precedes radiographic progression by up to 4 months and is associated with a measurable clinical decline. This work provides a viable window of therapeutic opportunity for future clinical trial design.

### Prospective Validation

The task to clearly define what qualifies as objective radiographic tumor response to therapy is ongoing. We are currently working to prospectively validate SI as part of a centrally reviewed, newly diagnosed GBM clinical trial (NRG-BN007). Prospective validation of this proposed imaging biomarker will be key to establishing signal intensity assessment in neuro-oncology (SANO) as an important tool for determining high-grade glioma response to therapy and expanding the lead-time for tumor treatment.

### Study Limitations

General study limitations were related to the retrospective study design restricting variables such as gathering patient reported outcomes and time of imaging and clinical follow-up. While using the NANO scale helped to standardize clinical assessments, these criteria have not been prospectively validated and do not comprehensively assess mood, quality-of-life, or other brain tumor symptoms that are central to GBM care ^52,53^. Finally, we did not fully explore sex-discrepant outcomes aside from noting the relative increased time to SI in young females. Future studies should seek to better discern gender- and sex-dependent outcomes.

## Data Availability

All data is available as noted below or stored as secured files within our institution and can be furnished upon request. A peer-reveiw for scientific journal publication is pending and updates will otherwise be published in the on-line or printed journal as well.

## CAPTIONS

**Supplemental Figure S1. (A)** Probability of outcomes based on landmark 6- and 12 month PFS and OS in patients with SI-pos vs. SI-neg status. **(B)** Two-by-two Contingency Tables based on pathology confirmation after repeat resection at 1^st^ progression (n=39 cases).

**Supplemental Figure S2**. Multivariate Analysis Model evaluating PFS and OS Hazard Ratios for SI after adjusting for age, sex, and KPS.

**Supplemental Figure S3. (A)** Decision Tree (Recursive Partitioning) determined factors the most influenced death vs survival. Female sex and age ≥64 as the next two most significant nodes impacting survival. **(B)** Graphic of PFS and OS (including best fit trendlines) in relationship to increasing average RC/V ratio. RC/V ratio (≥4, **dashed line**) was most significant determinant of death vs survival.

**Supplemental Figure S4**. Exploratory Data. Survival probability of SI-neg and SI-pos groups stratified by methylation status. **(Left K-M curve)** PFS. **(Right K-M curve)** OS. *Signal Intensity* (*SI*); *Methylated* (*M*); *Negative* (*neg*); *Positive* (*pos*); *Unmethylated* (*U*)

**Supplemental Figure S5**. Time to SI (TTSI) divided into three separate temporal periods demonstrating association with PFS and OS. **Early TTSI** < 5 months; **Intermediate TTSI** ≥5 but <11 months; and **Late TTSI** ≥11 months.

**Supplemental Figure S6**. Time from SI to progression (TTSI-P) stratified by MGMT methylation status. *Methylated* (*m*); *Unmethylated* (*u*); *Unknown* (*unk*)

**Supplemental Figure S7**. Univariate analysis of time from SI to progression (TTSI-P) stratified by age, MGMT methylation status, sex, KPS score at time of diagnosis, and MIB proliferation status.

**Supplemental Figure S8**. (A, Left image) Quantitative analysis of MR brain imaging for FLAIR SI within the vitreous fluid of the globes (eye) (left image, averaged two orbital measurements *O1,O2*) as compared to V (right image, white circles *d,e,f*). This signal difference is isointense. (A, Right image) Quantitative analysis of brain MR imaging for FLAIR SI within RC (averaged 3 measurements *a,b,c*). (B.) RC/E ratio measures compared to RC/V ratio measures for three timepoints: pre-SI, SI, and prog. *Eye* (*E*), *Progression* (*Prog*), *Resection cavity* (*RC*), *Ventricle* (*V*)

## Notes

### Competing Interest Statement

The authors have declared no competing interest.

### Funding Statement

FUNDING. Geisinger Foundation [01100000000000, Account #341020] via Philanthropic donations by Mr. Jeff Erdly, Mr. Jerry Sandel, The Lowe Family, and The Comp Family.

